# Method to Test Etiologic Heterogeneity Among Non-Competing Diagnoses that Accrue Over Time: Application to Discriminating the Impact of Perinatal Exposures on Autism and Attention Deficit Hyperactivity Disorder

**DOI:** 10.1101/2023.08.20.23294335

**Authors:** Amy E. Kalkbrenner, Cheng Zheng, Justin Yu, Tara E. Jenson, Thomas Kuhlwein, Christine Ladd-Acosta, Jakob Grove, Diana Schendel

## Abstract

**Background:** Testing etiologic heterogeneity – whether a disorder subtype is more or less impacted by a risk factor, is important toward understanding causal pathways and optimizing statistical power. The study of mental health disorders especially benefits from strategic sub-categorization because these disorders are heterogenous and frequently co-occur. Existing methods to quantify etiologic heterogeneity are not appropriate for non-competing events in an open cohort of variable-length follow-up. Thus, we developed a new method.

**Methods:** We estimated risks from urban residence, maternal smoking during pregnancy, and parental psychiatric history, with subtypes defined by the presence or absence of a co-diagnosis: autism alone, attention deficit hyperactivity disorder (ADHD) alone, and joint diagnoses of autism+ADHD. To calculate the risk of a single diagnosis (e.g. autism alone), we subtracted the risk for autism+ADHD from the risk for autism overall. We tested the equivalency of average risk ratios over time, using a Wald-type test and bootstrapped standard errors.

**Results:** Urban residence was most strongly linked with autism+ADHD and least with ADHD only; maternal smoking was associated with ADHD only but not autism only; and parental psychiatric history exhibited similar associations with all subgroups.

**Conclusions:** Our method allowed the calculation of appropriate p values to test strength of association, showing etiologic heterogeneity wherein 2 of these 3 risk factors exhibited different impacts across diagnostic subtypes. The method used all available data, avoided neurodevelopmental outcome misclassification, exhibited robust statistical precision, and is applicable to similar heterogeneous complex conditions using common diagnostic data with variable follow-up.

## Introduction

Etiologic heterogeneity is when subtypes of a disease or condition are influenced by different risk factors. The subtypes may be defined by level of severity, genetic markers, co-morbidities, or many other dimensions, and are supported in being causally distinct (e.g. etiologically heterogenous) when differentially influenced by one or a suite of causes. The study of etiologic heterogeneity, and valid approaches to do so, optimizes the goals of improving health, as follows. Observing that one subgroup is more (or less) impacted by a hypothesized causal factor, whether an environmental agent like tobacco smoke or a genetic variant, provides insight into shared versus distinct causal pathways. This insight can be combined with other knowledge, such as clinical and phenotypic dimensions of the subtype, and pathophysiological and toxicological information about the causal risk factor, to construct more complete maps of disease etiology. An additional advantage is enhanced statistical power, arising from isolating a more etiologically homogeneous subgroup that is more responsive to a given risk factor leading to a stronger measure of association, which would have been diluted had that subgroup been combined with non-responsive subgroups (with methodologic studies to demonstrate this potential).^1^ Lastly, better targeted interventions may follow from identifying distinct subtypes.

Mental health, psychiatric, and neurodevelopmental disorders especially benefit from studies of etiologic heterogeneity. Many of these conditions are highly heterogenous, for example autism, a diagnosis used for persons with a wide range of severity and functioning, which has led to the hypothesis that autism is an aggregation of several underlying disorders with distinct pathophysiologies. The search for more homogenous subtypes or new classifications to advance the understanding and treatment of mental health disorders is evidenced by searches for endophenotypes,^2^ efforts to sub-classify based on genetic markers,^3^ and the RDoC initiative of the National Institute of Mental Health.^4^

One simple, but common, means of subtyping and evaluating etiologic heterogeneity uses co-diagnosis with another mental health disorder. Here we will focus on the co-diagnosis of autism with attention deficit hyperactivity disorder (ADHD), which is common: 20-50% of children with ADHD also meet diagnostic criteria for autism and 30-80% of autistic children also meet diagnostic criteria for ADHD.^5 6 7,8^ This co-occurrence even has an unofficial term in social use created by combining both words: “AuDHD”.

Example publications that have examined etiologic heterogeneity with autism and a co-diagnosis have concluded that: maternal smoking in pregnancy posed a higher risk for autism without co-occurring intellectual disabilities;^9^ prenatal acetaminophen exposure posed a higher risk for autism with ADHD-like symptoms;^10^ prenatal anti-depressant exposure posed a higher risk for autism without co-occurring intellectual disabilities;^11^ and maternal metabolic conditions posed a higher risk for autism with co-occurring intellectual disabilities.^12^ Common to these studies is drawing conclusions by comparing the magnitude of the measure of association, that is, equivalence of the risk ratios, without explicit regard for confidence interval overlap. The drawback to this approach is that differences may be the result of random error. A direct and appropriate statistical test would generate stronger conclusions.

While a body of literature has developed methodologies to test etiologic heterogeneity while considering random error, much of this pertains to “either-or” health outcomes that cannot occur together (fully competing events), such as cancer subtypes.^13–15^ Etiologic heterogeneity can also be evaluated for health outcomes that can co-occur, as is often the focus for mental health disorders and our example here, that is: non-competing events, also called comorbidities or co-diagnoses.^16,17^ Existing statistical methods to evaluate non-competing subtypes assume that all persons are followed for the same time or that time information is not available, as reviewed by Zabor and Begg.^18^

Allowing for the accrual of diagnoses over time when comparing disorder subtypes is important, because a snapshot from a single time point will for some misclassify the eventual status. The proportion who change diagnostic status can be substantial, as evidenced by a lack of a plateau for autism diagnoses even into adulthood.^19^ Given that diagnostic criteria for autism (e.g. the Diagnostic and Statistical Manual) require childhood onset of symptoms, and preponderance of evidence of early life etiology, we assume that the final phenotypic subtype was present early, even while the clinical recognition may only become apparent with age. To date, published methods are insufficient for this type of etiologic heterogeneity testing where co-diagnoses accrue over time. In our investigation, subtypes are defined by joint or separate diagnoses of autism and ADHD to yield 4 combinatory statuses: neurotypical (not diagnosed with autism or ADHD), autism only, ADHD only, and autism+ADHD. These non-competing events might be marginally modeled by a Cox model. Some of these diagnostic subgroups are only partially observable, however, because without complete follow-up, we cannot definitively discern the autism only or the ADHD only subgroups (i.e., with additional follow-up time an individual classified as autism only may convert to autism+ADHD). To analyze this type of data, a multi-state model could be used if the change from autism only to autism+ADHD is the progression of interest (e.g., in a study of different referral and diagnosis practices). More often the question of interest is about causality (e.g. etiologic heterogeneity) among the subtypes.

Therefore, we have developed a new approach to statistically test etiologic heterogeneity for non-competing events that accrue over time. As proof of principle, we made quantitative comparisons of the strength of associations between 3 putative perinatal risk factors for autism and ADHD (urban residence at birth, maternal smoking during pregnancy, and parental previous psychiatric diagnoses), with three neurodevelopmental phenotypic subgroups (autism only, ADHD only, and autism+ADHD).

## Methods

### Included Persons

We included persons from iPSYCH, a population-based Danish case-cohort study of several psychiatric disorders,^20^ which includes singletons with known mothers who were alive and resided in Denmark on their first birthday. We included all persons with a diagnosis of autism or ADHD, and, to represent the cohort base, a random sample of approximately 2 percent of eligible persons, without regard to psychiatric diagnostic status, born January 1, 1991 to December 31, 2005, followed until emigration, death, diagnosis, or the end of follow-up (December 31, 2012). Variables were largely derived from Danish registries, linked using personal identification numbers.

### Psychiatric Diagnoses of Index Persons and Parents

Information on the diagnosis of autism (ICD-10 diagnosis of F84.0, F84.1, F84.5, F84.8 or F84.9) or ADHD (F90.0) were from the Danish Psychiatric Central Research Register (PCRR) occurring at age 1 year or later.^21^ Follow-up time was calculated as the date of birth until the diagnosis or diagnoses. Parental history of mental disorders were derived from the PCRR based on any reported ICD 8 (290-315) or ICD 10 (F00-F99) diagnosis for either the mother or father before the child’s birth.

### Covariates

Sex, date of birth, parental age, maternal marital status, and parental immigrant status were obtained via the Danish Civil Registration System.^22^ Parity and interpregnancy interval were obtained via the Danish Medical Birth Register.^23^ Parental income was obtained using the Registers on Personal Income and Transfer Payments.^24^ Parental educational attainment was obtained using the Danish Education Registers.^25^ Father’s educational attainment and employment pertain to the index’s date of birth; mother’s educational attainment and employment pertain to two years prior to index’s date of birth to reflect typical occupation not potentially impacted by pregnancy and maternity leave.

### Measurement of Urbanicity

Information on urbanicity was defined based on maternal address at the time of the child’s birth, from the Danish Civil Registration System.^22^ Briefly, the system records the address for all persons in Denmark and assigns a municipality code.^26^ The municipality codes are converted to a 12-category measure of urbanicity, which describes the type and size of the city or town,^27^ a method that captures important health gradients in Denmark, but is distinct from population density. For analysis, we compared the most urban region (defined as residence in the capital region, Copenhagen) to the most rural region (defined as residence in other municipalities less than 50 percent urban).

### Measurement of Maternal Smoking in Pregnancy

Data on maternal smoking during pregnancy, reported to the midwife at the first prenatal visit, was from the Danish Medical Birth Register.^23^ For analyses, we compared women who ever smoked in pregnancy (who stated that they were current smokers or reported stopping smoking during the first trimester) versus women who stated that they had never smoked in this pregnancy.

### Statistical Analysis: Multiple Imputation to Account for Potential Selection Bias

About 13% of eligible persons were missing information on one or more variables and would have been excluded from a complete-case analysis. Having a missing variable was slightly less common for persons with an ADHD diagnosis (eTable 1) than an autism diagnosis. Persons with missing data were substantially more likely to be immigrants, born in earlier years, and have unmarried parents (eTable 2). Given the possibility of inducing a selection bias, we elected to predict and fill in missing data using multiple imputation via chained equations (MICE) using all covariates and respective diagnosis follow-up times as predictors.^28^ We separately imputed for persons diagnosed with autism only, ADHD only, autism and ADHD, and without a diagnosis, then recombined the datasets for analysis.

### Statistical Analysis: Estimating Risk and Risk Ratios

We used Cox regression models to predict the individual potential risk of receiving a diagnosis of autism, ADHD, or both, for 20 equal time periods starting at 1 year of age and for the entire duration of follow-up. Given the case-cohort sampling frame, we used inverse probability weighting to account for the fact that the sub-cohort represented 2 percent of the included Danish population.^29^

We adjusted for several variables because they were associated with the exposure and neurodevelopmental outcome or because they may improve statistical precision (e.g. child sex). For continuous confounders (e.g. maternal age), we selected either the original variable coding, a log transform, a quadratic transform, or categorical coding that had the lowest Akaike Information Criterion (AIC). We adjusted for the following covariates in all models: child sex, birth year (categorical), inter-pregnancy interval (categorical), maternal marital status, maternal and paternal educational attainment (categorical), income (quadratic), employment status (categorical), age (quadratic), immigrant status, along with the risk factors of interest: parental psychiatric diagnosis, urbanicity, and maternal smoking.

The key innovation of our method is our approach to calculating risk for unobservable subgroups: those with a single diagnosis. Rather than using a person’s final diagnostic status at the end of follow-up, which reflects the status at an arbitrary point of time that may change with more follow-up, we used a “subtraction” method. To obtain the risk of being diagnosed only with autism, we relied on the fact that the risk of being diagnosed with autism is the sum of being diagnosed with autism without ADHD (autism only) added to the risk of a co-diagnosis (autism+ADHD). We subtracted the risk of co-diagnosis from the risk of being diagnosed with autism overall at each time point. We similarly performed this calculation for ADHD. The estimated risks of autism only and ADHD only are not guaranteed to be positive; however, since the estimator is consistent, the probability of being negative will converge to 0 when the sample size goes to infinity. In our application, we did not see negative estimated risks. The population average potential risks were estimated as the inverse probability weighted mean of the individual potential risks among the cohort. The time dependent risk ratios (RR) comparing high to low exposure groups were computed over the follow-up period and then the average risk ratios were computed as a summary for the effect of that exposure.

The results from 20 imputed datasets were averaged to provide the final estimations. We obtained confidence intervals using a normal approximation with standard errors estimated via Bayesian bootstrapping with 100 iterations.^30^

### Statistical Analysis: Testing Etiologic Heterogeneity

Adapting the idea of quantifying heterogeneity by a ratio of association measures, that is, ratio of risk ratios (RRR),^31^ we tested whether the RRR averaged over follow-up time equaled 1 for all pairs of subtypes (e.g. autism only vs. ADHD only), using a Wald type test with a pre-specified contrast matrix. Unlike previous studies which inferred etiologic heterogeneity based only on a regression coefficient or, equivalently, a conditional relative risk, here we considered the population average relative risk as a target parameter (a causal parameter that does not depend on model specification) and thus has better causal interpretation and is more robust to model misspecification.^32^ A bootstrapped variance-covariance matrix was computed to handle the correlation between parameters for different outcomes. The estimated survival functions for autism+ADHD, ADHD only, and autism only for each group are consistent and asymptotically multivariate normally distributed when the model is correctly specified, and thus the RRR calculated from these survival functions will also be consistent and normally distributed, which guarantees that the Wald-type test using bootstrapped standard errors had a valid test size.

### Statistical Analysis: Conventional Alternate Analyses

We performed two conventional alternative analyses to directly compare to our “subtraction” approach for the urban residence exposure. From our data we constructed 4 closed cohorts of different follow-up lengths. We started with a cohort followed from birth to age 6 years (school entry) and then proceeded by 4-year intervals with different lengths of follow-up until 18 years. For each closed cohort, we calculated odds ratios for each diagnostic subgroup, defined at the end of the follow-up period, adjusting for all variables in our primary models.

Secondly, we modeled odds ratios for the entire sample (open cohort) for single and co-diagnoses assigned at the end of the individual’s follow-up (which ranged from 7 to 22 years). We used the confidence limit ratio (upper 95% confidence interval divided by the lower 95% confidence interval) to measure precision around these measures of association.

## Results

### Population and Diagnostic Characteristics

We included 42,831 persons who were part of the sampled sub cohort and/or had a diagnosis of autism (10,278), ADHD (11,131), or both (2,686), within our follow-up time. The average follow-up duration and standard deviation in years was 9.6 (4.3) for persons diagnosed with autism only, 11.2 (4.2) for persons diagnosed with ADHD only, and 10.1 (3.5) for persons diagnosed with autism and ADHD. In general, persons with an autism or ADHD diagnosis were more likely to be male, to have non-immigrant parents, and to have a parent diagnosed with a psychiatric disorder (Table 1). Some demographic patterns differed across the diagnostic subgroups, for example, persons with ADHD (but not autism) were more likely to have non-married parents; persons with autism (but not ADHD) were more likely to have mothers or fathers in the oldest age group; and persons with ADHD (but not autism) were more likely to have parents with lower education levels. About a quarter (23.3%) of mothers smoked in pregnancy during the study years, with higher proportions of mothers smoking in pregnancy for persons diagnosed with autism (26.8%) and ADHD (41.7%). Overall, 14.7% of mothers in the sample resided in the most urban (capital) region at birth, with a higher proportion among persons diagnosed with autism (19.2%) and a lower proportion among persons diagnosed with ADHD (11.6%).

**Table 1.**
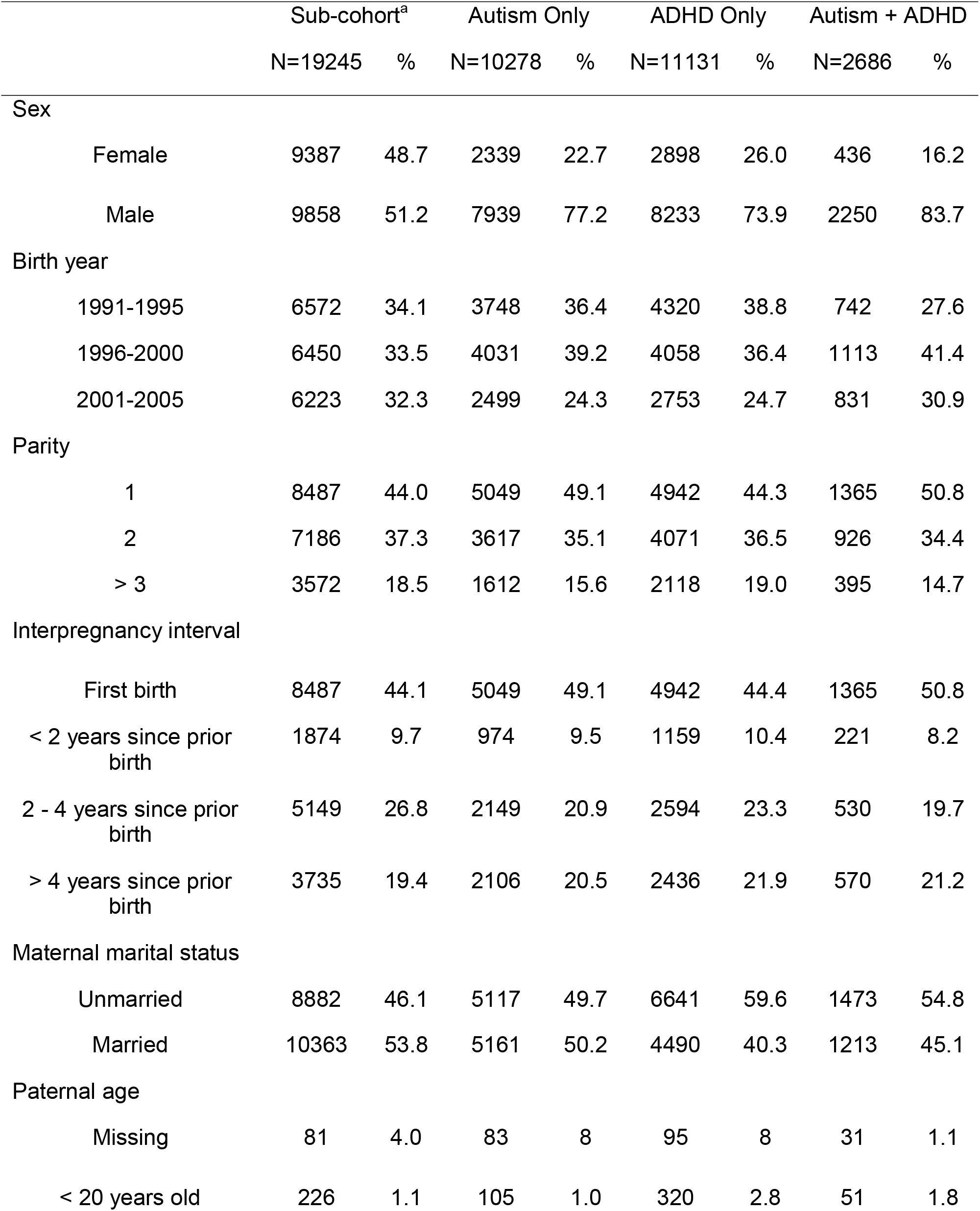
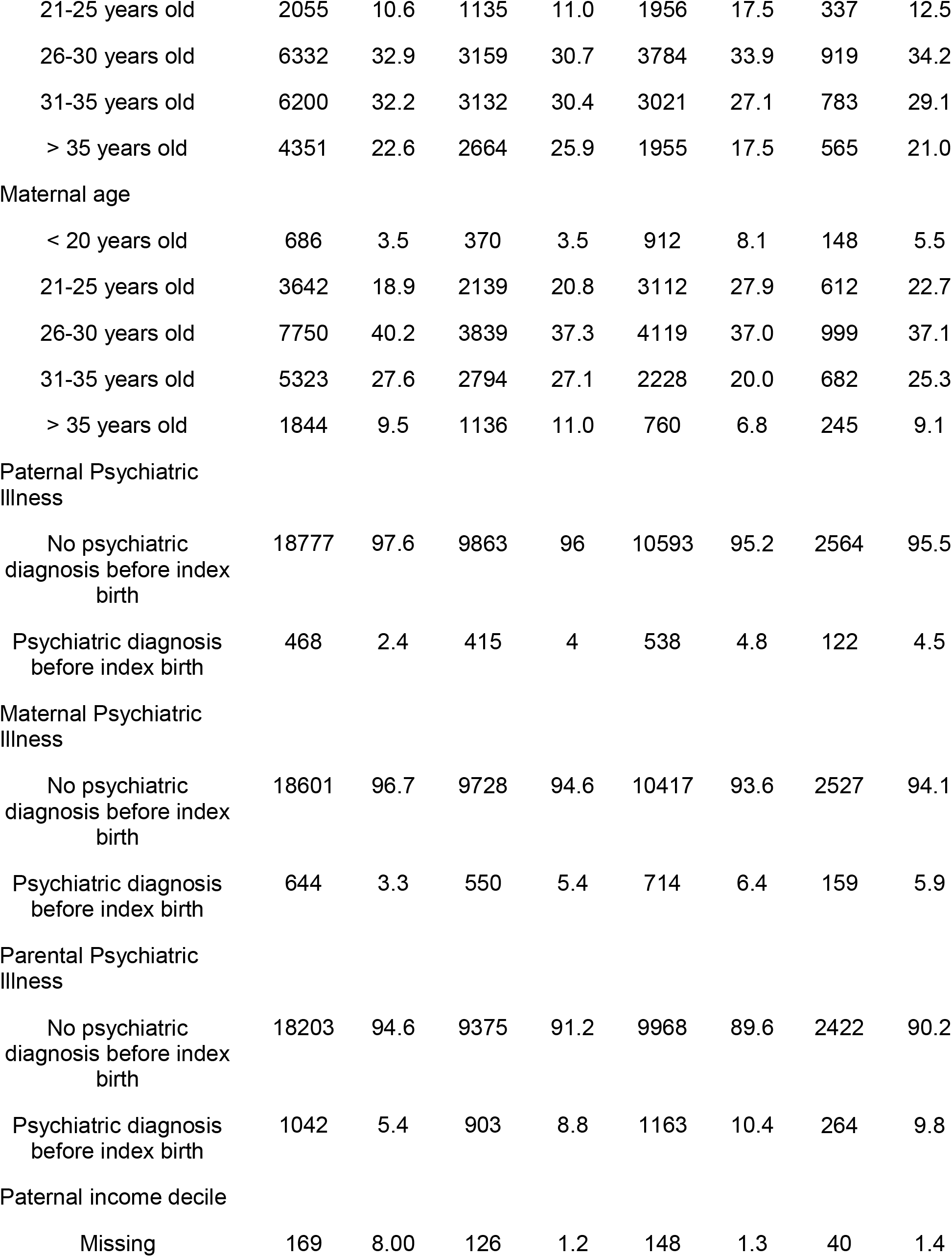
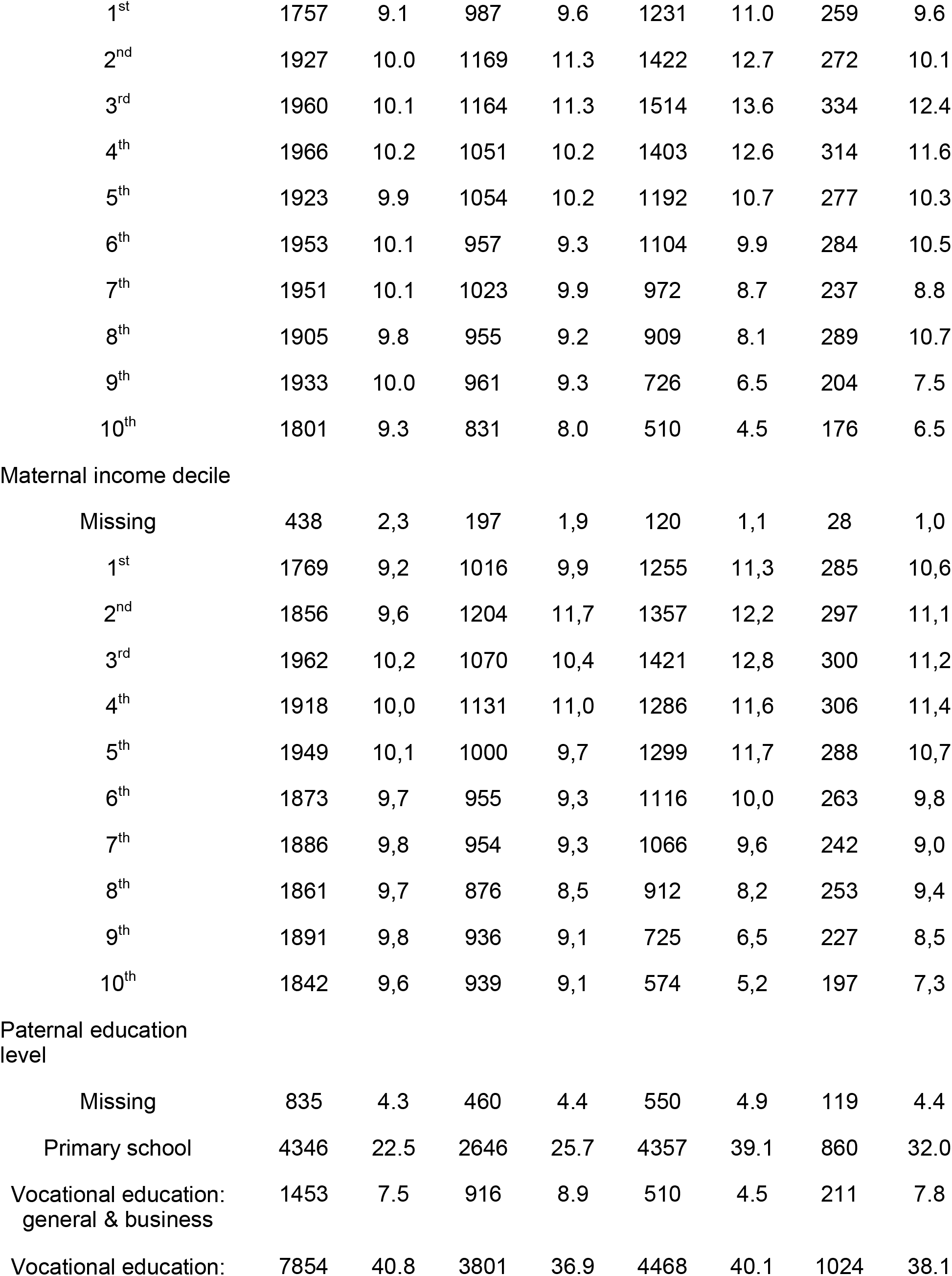
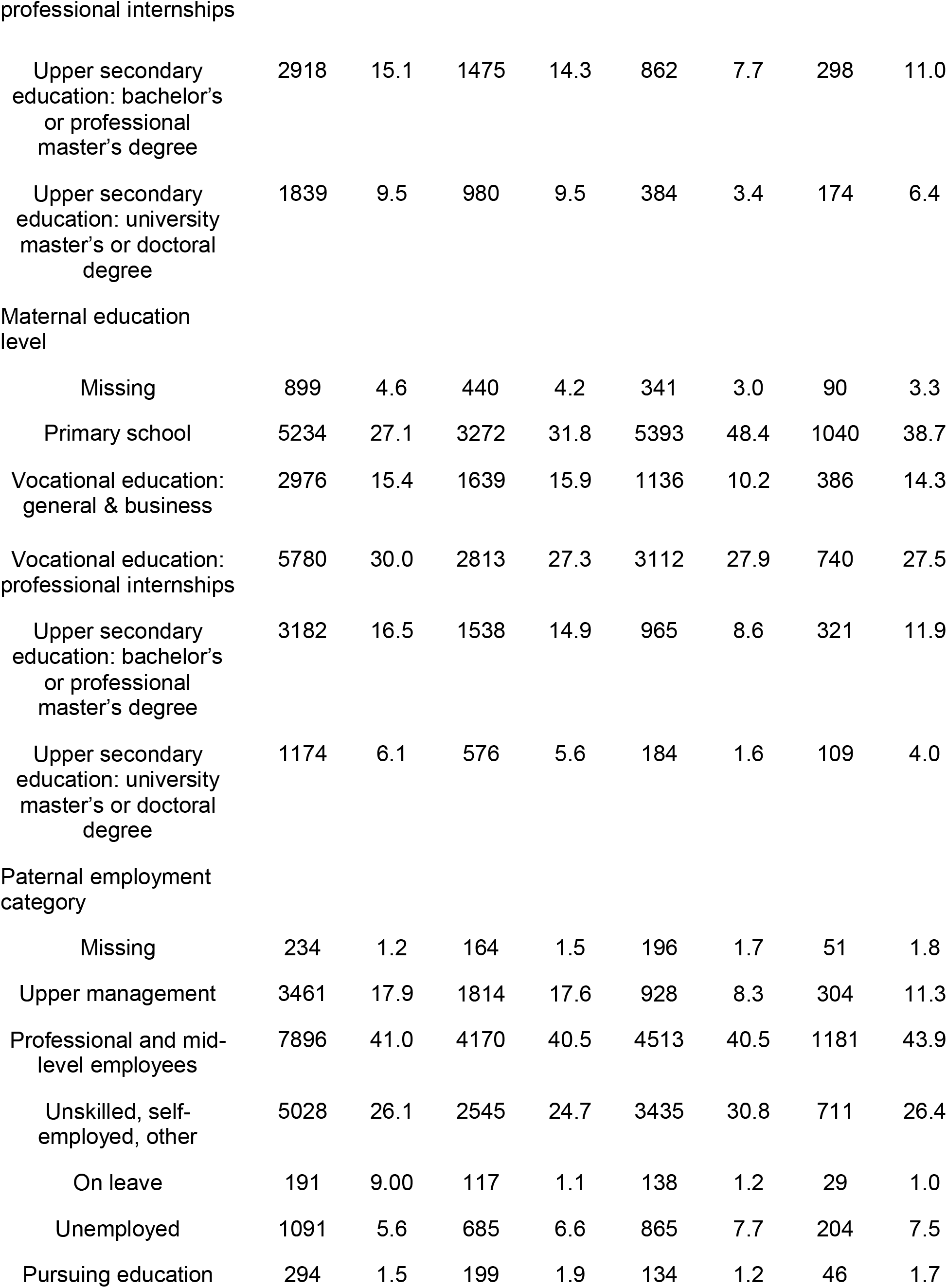
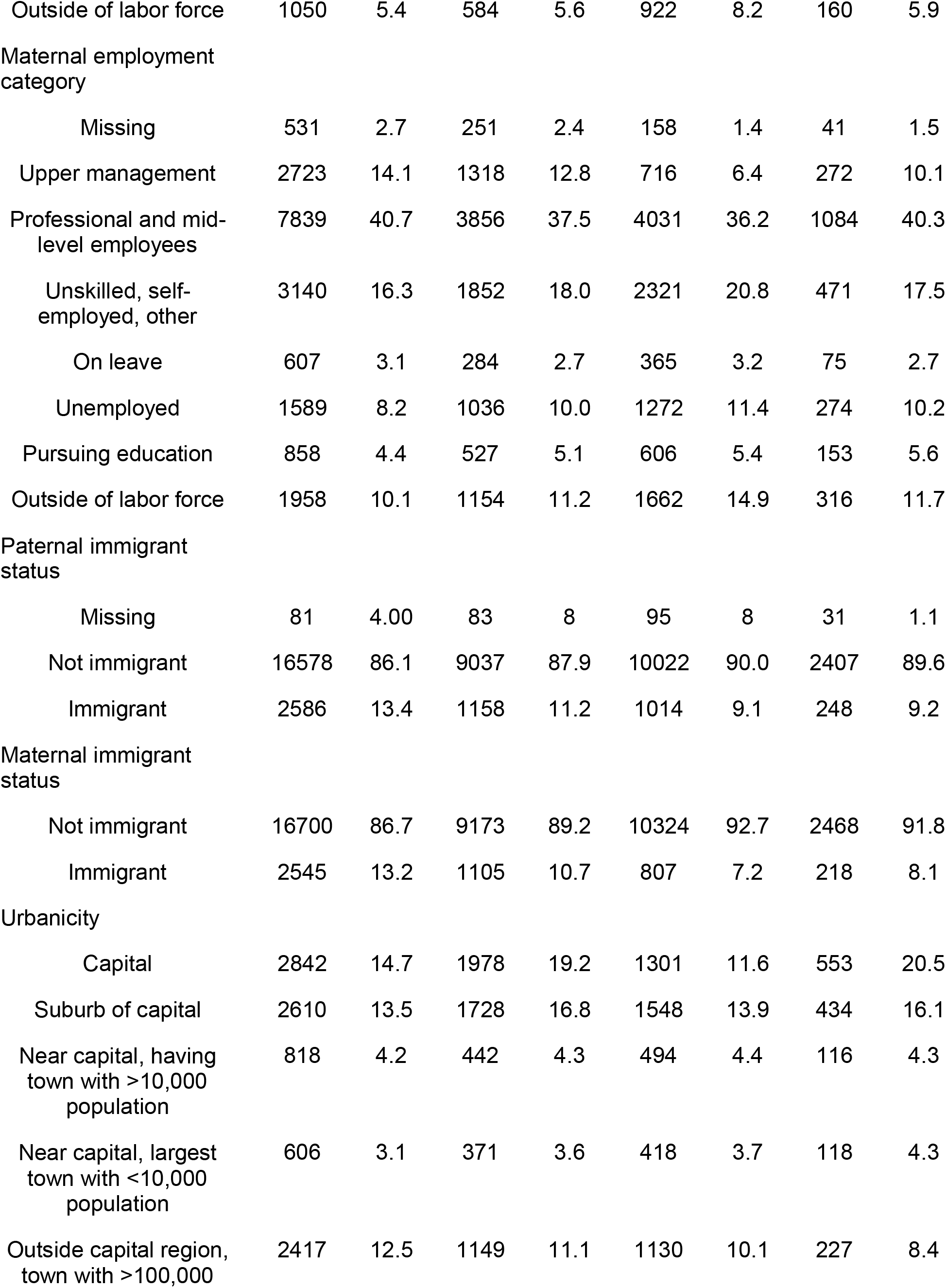
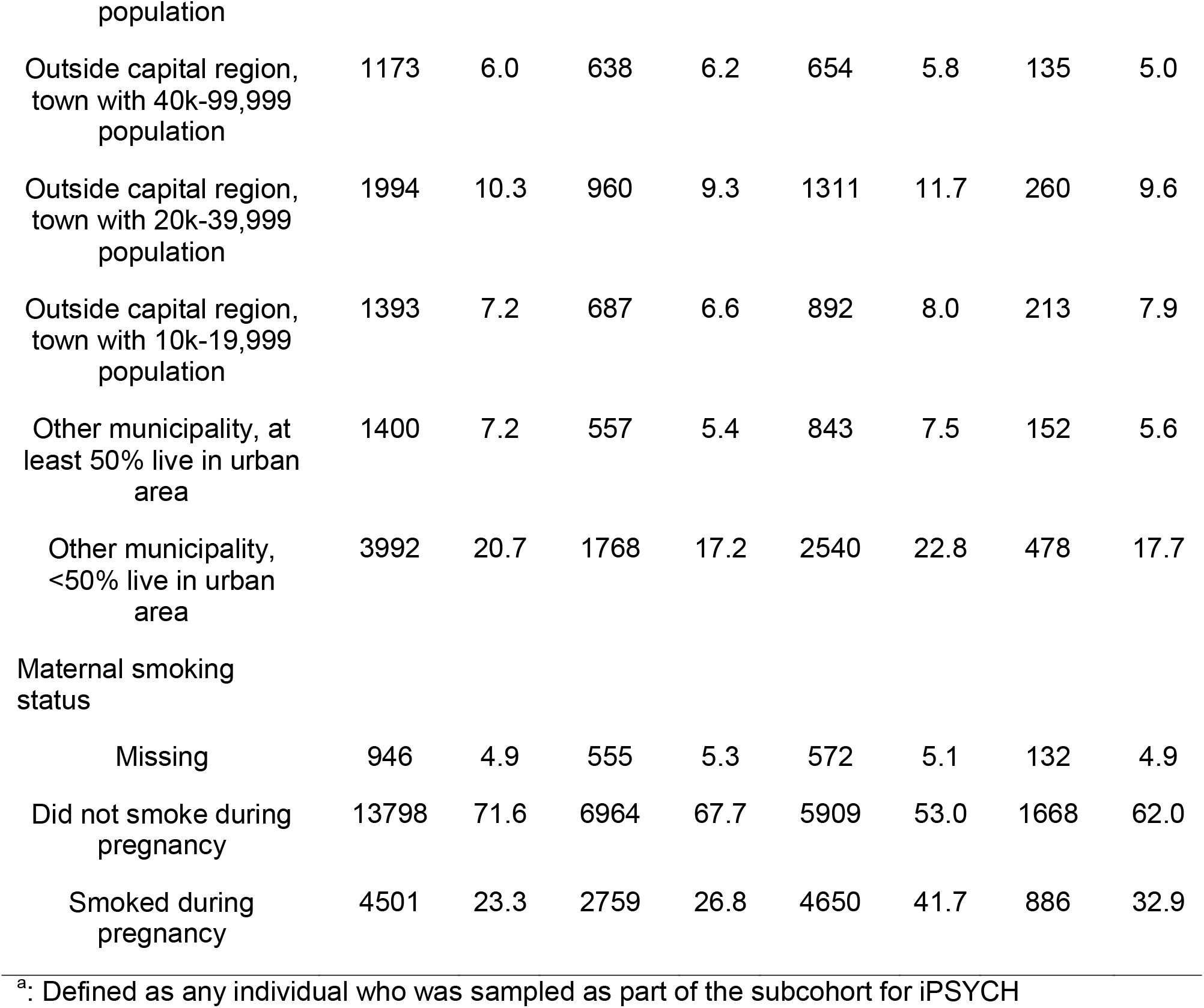
Characteristics of Persons in iPSYCH Study Born Between 1991 and 2005 by Diagnostic Status at End of Follow-up.

### Urbanicity Associations

After adjusting for confounders, being born in the most urban environment compared to the most rural environment was associated with a small increased risk for ADHD only: (red in Figure 1) and average risk ratio over time (RR) = 1.2 (Table 2), a larger increased risk for autism only: (blue in Figure 1) and RR = 1.7 and a highest risk associated with being co-diagnosed with autism+ADHD: (purple in Figure 1) and RR = 2.2. Our subtraction method to compare whether these measures of association were statistically distinct confirmed that urbanicity exerted a stronger influence on autism+ADHD vs. autism only (p = 0.0007) (Table 3). The lower association with ADHD only (RR = 1.2) was statistically distinct from that for autism only and autism+ADHD (both p < 0.0001).

**Figure 1.**
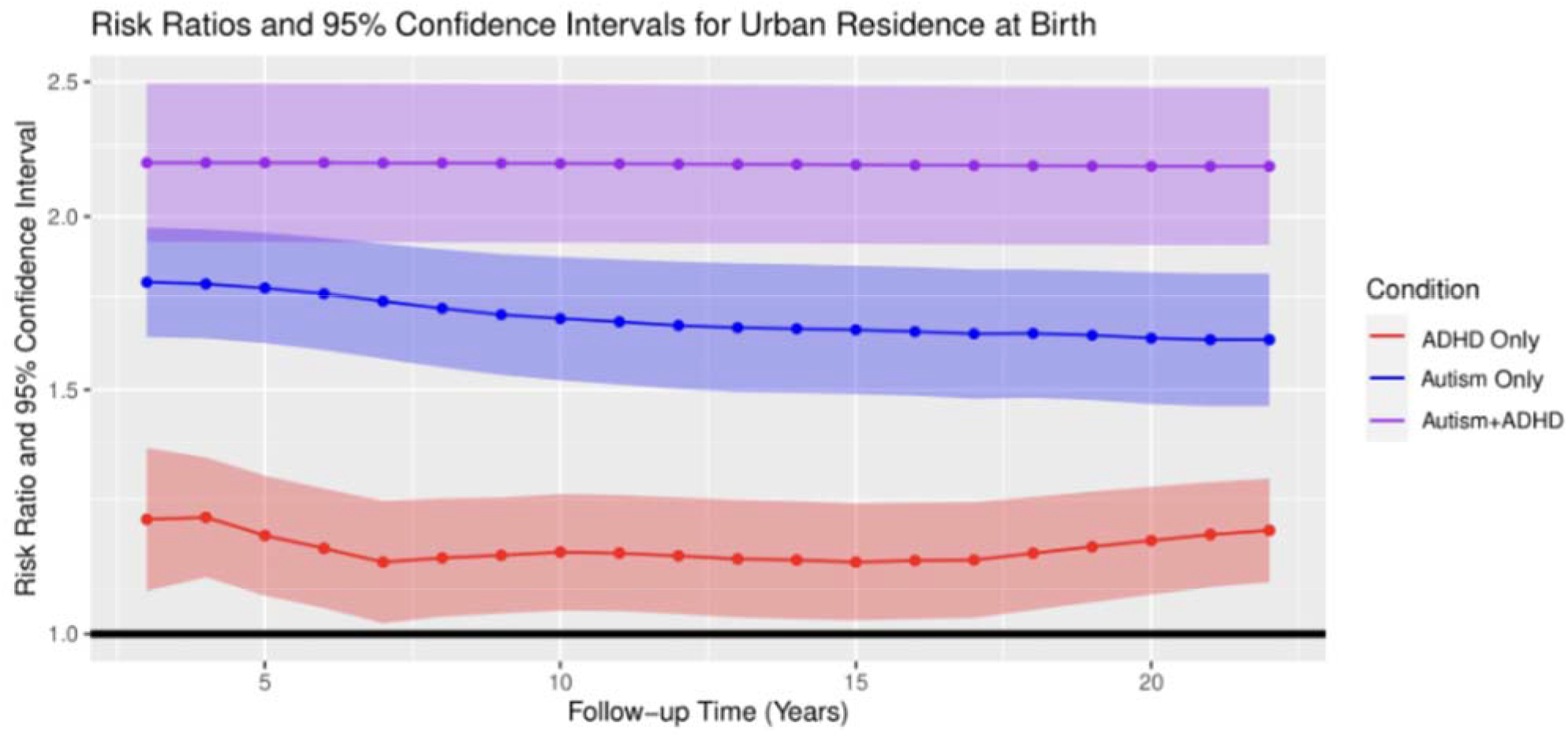
Comparison of Adjusted Risk Ratios Over Time for Urban Residence on Autism and ADHD Diagnostic Subgroups. ^a^: Models were adjusted for sex, birth year, interpregnancy interval, maternal marital status, maternal and paternal age (quadratic term included), maternal and paternal psychiatric diagnosis prior to index birth, maternal and paternal income deciles (quadratic term included), maternal and paternal educational attainment, maternal and paternal employment status, maternal and paternal immigrant status, and maternal smoking ^b^: Confidence intervals (CI) were determined using 100 bootstrap iterations ^c^: Comparing most urban category (Capital region) to most rural category (Other region with <50% living in urban area.

**Table 2:** Average Adjusted Risk Ratios for Urban Residence, Maternal Smoking, and Parental Psychiatric Diagnosis for Autism and ADHD Diagnostic Subgroups, iPSYCH, Born Between 1991 and 2005, Denmark.

|  | Autism<br>only<sup>a</sup> | 95% CI<sup>b</sup> | ADHD<br>only<sup>a</sup> | 95% CI<sup>b</sup> | Autism +<br>ADHD<sup>a</sup> | 95% CI<sup>b</sup> |
| --- | --- | --- | --- | --- | --- | --- |
| Urban<br>Residence<sup>c</sup> | 1.7 | 1.5, 1.9 | 1.2 | 1.1, 1.3 | 2.2 | 1.9, 2.5 |
| Maternal<br>Smoking | 1.1 | 1.0, 1.1 | 1.5 | 1.4, 1.6 | 1.2 | 1.1, 1.4 |
| Parental<br>Psychiatric<br>Diagnosis | 1.5 | 1.3, 1.6 | 1.4 | 1.3, 1.6 | 1.4 | 1.2, 1.6 |

<sup>a</sup>: Models were adjusted for sex, birth year, interpregnancy interval, maternal marital status, maternal and paternal age (quadratic term included), maternal and paternal psychiatric diagnosis prior to index birth, maternal and paternal income deciles (quadratic term included), maternal and paternal educational attainment, maternal and paternal employment status, maternal and paternal immigrant status, urbanicity, and smoking
<sup>b</sup>: Confidence intervals (CI) were determined using 100 bootstrap iterations
<sup>c</sup>: Comparing most urban category (Capital region) to most rural category (Region with <50% living in urban area)

**Table 3:** P Values Testing Equivalency of Average Adjusted Risk Ratios^a, b^ Over Time for Urban Residence, Maternal Smoking, and Parental Psychiatric Diagnosis with Autism and ADHD Diagnostic Subgroups, iPSYCH, Born Between 1991 and 2005, Denmark.

|  | Autism Only vs<br>ADHD Only | Autism Only vs<br>Autism+ADHD | ADHD Only vs<br>Autism+ADHD |
| --- | --- | --- | --- |
| Urban Residence<sup>c</sup> | < 0.0001 | 0.0007 | < 0.0001 |
| Maternal Smoking | < 0.0001 | 0.0016 | 0.0004 |
| Parental Psychiatric Diagnosis | 0.4610 | 0.4582 | 0.8199 |

<sup>a</sup>. P-values were obtained from Wald test with degree of freedom 1 using variance-covariance matrix computed from 100 Bootstrap samples.
<sup>b</sup>. Models were adjusted for sex, birth year, interpregnancy interval, maternal marital status, maternal and paternal age (quadratic term included), maternal and paternal psychiatric diagnosis prior to index birth, maternal and paternal income deciles (quadratic term included), maternal and paternal educational attainment, maternal and paternal employment status, maternal and paternal immigrant status, urbanicity, and smoking
<sup>c</sup>: Comparing most urban category (Capital region) to most rural category (Region with <50% living in urban area)

### Smoking Associations

Maternal smoking in pregnancy was associated with ADHD only (red in Figure 2) average RR over time = 1.5 (Table 2) to a lesser extent with autism+ADHD (purple in Figure 2) RR = 1.2, and was not associated with autism only (RR = 1.1) (blue in Figure 2). All three of these measures of association were statistically distinct (Table 3).

**Figure 2.**
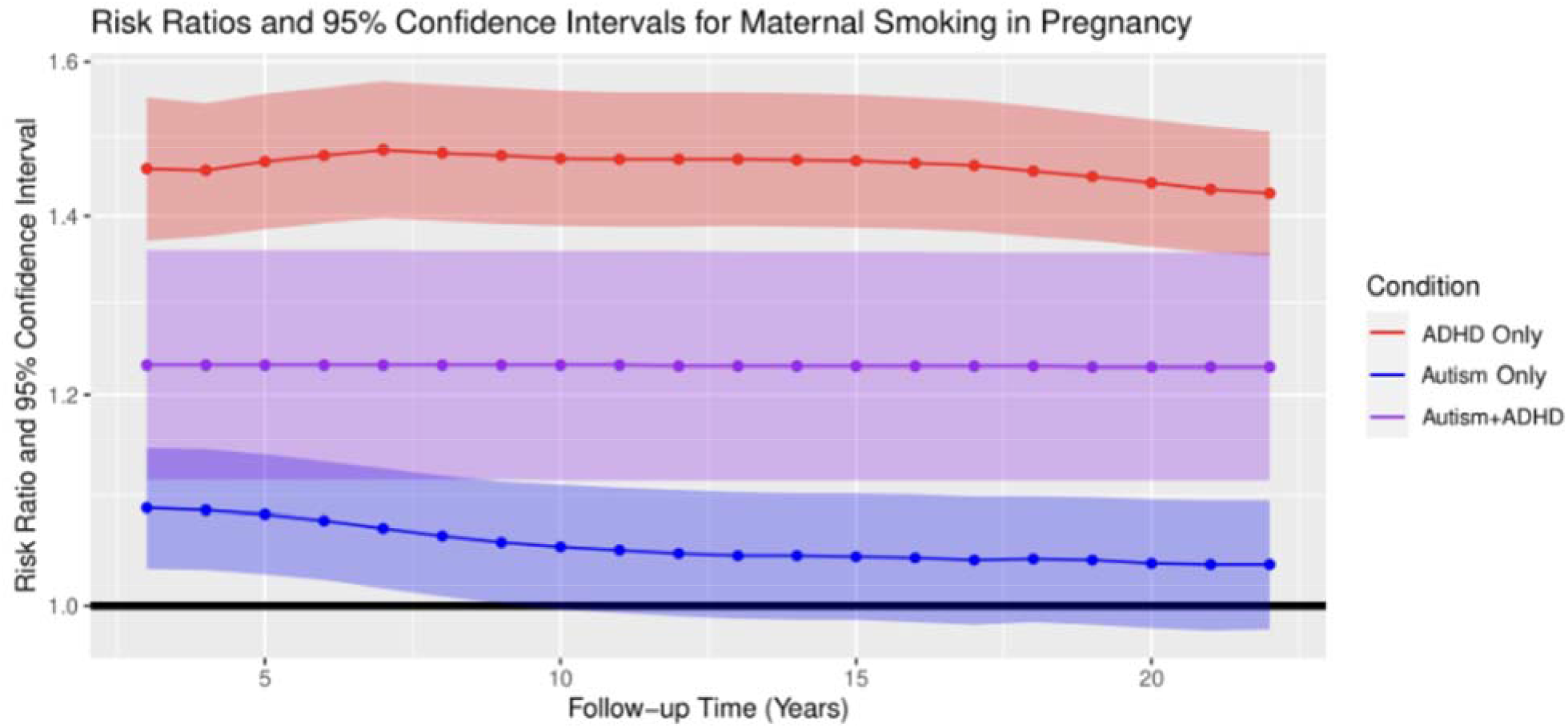
Comparison of Adjusted Risk Ratios Over Time for Maternal Smoking in Pregnancy on Autism and ADHD Diagnostic Subgroups. ^a^: Models were adjusted for sex, birth year, interpregnancy interval, maternal marital status, maternal and paternal age (quadratic term included), maternal and paternal psychiatric diagnosis prior to index birth, maternal and paternal income deciles (quadratic term included), maternal and paternal educational attainment, maternal and paternal employment status, maternal and paternal immigrant status, and maternal smoking ^b^: Confidence intervals (CI) were determined using 100 bootstrap iterations

### Parental Psychiatric Diagnoses Associations

Parental psychiatric diagnosis before the index person’s birth was associated with all three diagnostic subgroups with similar RRs (1.4 - 1.5) (Table 2 and Figure 3) and the measures of association were not statistically distinct (p values ranging from 0.4-0.8) (Table 3).

**Figure 3.**
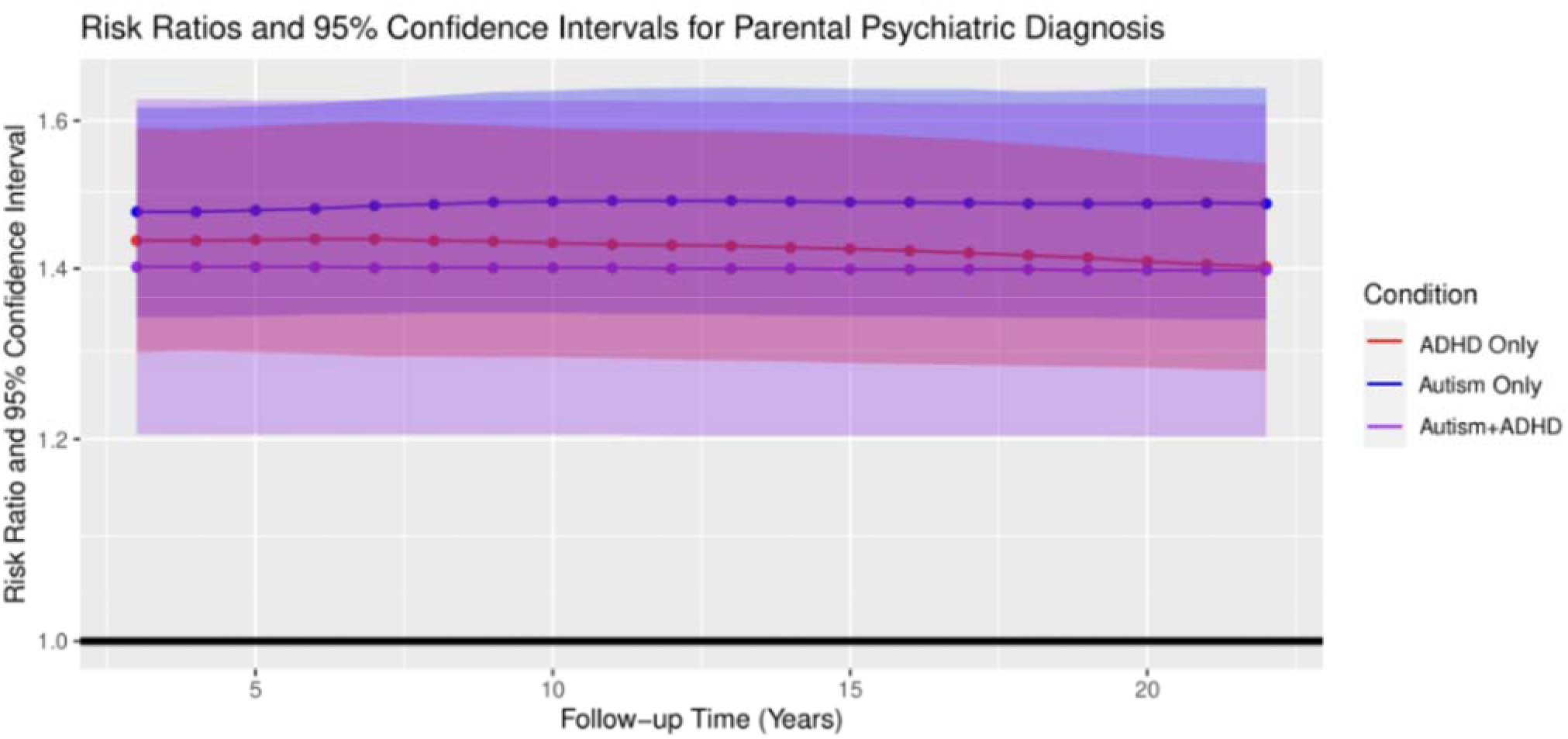
Comparison of Adjusted Risk Ratios Over Time for Parental Psychiatric Diagnosis on Autism and ADHD Diagnostic Subgroups. ^a^: Models were adjusted for sex, birth year, interpregnancy interval, maternal marital status, maternal and paternal age (quadratic term included), maternal and paternal psychiatric diagnosis prior to index birth, maternal and paternal income deciles (quadratic term included), maternal and paternal educational attainment, maternal and paternal employment status, maternal and paternal immigrant status, and maternal smoking ^b^: Confidence intervals (CI) were determined using 100 bootstrap iterations

### Conventional Alternate Analyses

Our 4 closed cohorts with different sample sizes and lengths of follow-up exhibited a pattern of higher prevalence with more follow-up for all three diagnostic subgroups, reflecting the ongoing diagnosis of both autism and ADHD with age (eTable 3). Notable proportions (3-8%) transitioned from single to co-diagnosis upon having longer follow-up in each period, and up to 21% of persons classified as co-diagnosis had a single diagnosis in a previous period (eTable 4).

When examining the impact of urban residence at birth, the direction of results were the same for these alternate analyses as compared to our new subtraction method (eTable 5). That is, urbanicity exerted the strongest impact on the co-diagnosis autism+ADHD, a moderate impact on being diagnosed with autism only, and the smallest impact on ADHD only. Precision was superior for our subtraction method compared to the closed cohorts, and this enhanced precision was especially notable for the more rare joint autism+ADHD subgroup, where the confidence limit ratio (CLR) was 1.3 vs. conventional methods with CLR ranging from 1.6-2.2. The open cohort approach yielded results that were strikingly similar in terms of magnitude and precision of measures of associations in comparison to our “subtraction” method. Importantly, none of the conventional approaches resulted in direct tests of etiologic heterogeneity, such as a p value testing equivalency of risk ratios.

## Discussion

We developed and applied a method to evaluate whether a risk factor (presumptive causal influence) exhibited a statistically distinct strength of association across different disorder subtypes, in a situation where non-competing events accrued over time. Our method tested whether the single diagnosis subgroups (autism only or ADHD only) and the autism+ADHD subgroup each potentially represented a distinct phenotype as evidenced by being influenced to a different degree by well-studied perinatal risk factors for neurodevelopment. By constructing an appropriate p value directly evaluating whether a given risk factor exhibited different strength of association, we can more confidently evaluate whether causal pathways to autism are independent of those for ADHD, and whether the co-diagnosis of autism+ADHD is distinct in causal influences from either single diagnosis, or moreso resembles autism or ADHD.

This method exhibits several strengths: It was not reliant on the arbitrary end of follow-up that often occurs in real world research settings, such as in the use of administrative datasets, which can lead to outcome misclassification for diagnoses which accrue with age. In fact, we confirmed a meaningful degree of misclassification in our constructed closed cohorts, where persons with only one diagnosis would later accrue a second diagnosis. Importantly, our method allowed the inclusion of the entire administrative dataset, despite variation in length of follow-up across the sample, and yielded increased precision compared to closed cohort methods. Our method is less sensitive to the parametric model form and is less computationally complicated compared to a bivariate survival model with cure proportion, another approach used to evaluate etiologic heterogeneity.^33^ Importantly, we were able to generate a p value directly comparing the equivalency of risk ratios to appropriately quantify the difference in risk factor strength. While a formal test of method validity is outside the scope here, the underlying normal distribution and consistency of the underlying parameters, along with the use of a bootstrapped standard error, implies that the resultant Wald test will have a valid test size. With enhanced precision and valid p values, more robust conclusions can be drawn regarding risk profiles that are distinct versus similar between non-competing outcomes than comparing the magnitude of RRs or by examining confidence interval overlap, as is frequently done in etiologic studies of neurodevelopment.

Residence in an urban area (urbanicity) has often been linked with a higher prevalence of psychiatric and neurodevelopmental conditions. Our approach yielded a clear picture that autism only and the co-diagnosis of autism+ADHD were linked with urban residence, whereas diagnosis with ADHD only was not. This pattern could indicate that causal factors in urban areas (environmental pollutants, crowding, noise, and other factors) may exert more influence in neurodevelopmental pathways leading to autism symptoms more so than ADHD symptoms. Another potential explanation is that receiving an autism diagnosis is more sensitive to awareness or access to medical specialists that cluster in urban areas compared to ADHD. Our method additionally demonstrated that the influence of urbanicity was stronger for co-diagnosis of autism+ADHD than autism alone. This observation is consistent with the hypothesis that urban residence is associated with an increased likelihood of receiving a neurodevelopmental diagnosis, generally, given that these persons received two diagnoses.

Maternal smoking in pregnancy has often been linked with suboptimal fetal development, presumably via toxic aspects of tobacco smoke influencing oxidative stress or impaired placental blood flow. We showed that maternal smoking links with ADHD alone were much stronger than with autism alone, with the autism+ADHD subgroup being intermediate in risk. These findings suggest that maternal smoking in pregnancy is more likely to lead to the ADHD phenotype, without influence on the autism phenotype. However, we note that we only used conventional adjustment for measured confounders here, which may insufficiently rule out complex confounding that is better addressed by family-based designs to assess whether influences of maternal smoking in pregnancy on neurodevelopment are truly causal.^34,35^

In contrast to urbanicity and maternal smoking, parental diagnosis with a psychiatric condition exhibited similar strengths of association for all three diagnostic subgroups that were not statistically distinct. This result suggests that the pathways linking parental psychiatric history to these subgroups are similar, and is consistent with findings in other studies.^36–39^ That the association may reflect shared etiologic mechanisms is also consistent with reports of considerable genetic overlap across autism, ADHD, and other psychiatric conditions.^40–42^ An alternate explanation is that this parental psychiatric diagnosis variable was non-specific, given that it combined across psychiatric diagnoses. The fact that our method did not discriminate differences for parental psychiatric history, although it did for urbanicity and maternal smoking, points to its utility in providing insight into the distinctiveness of risk factor profiles across potentially overlapping phenotypes.

Limitations of these findings include the reliance on documented diagnoses, which may have some degree of missed diagnosis. However, universal access to health care in Denmark reduces the chances for underdiagnosis of neurodevelopmental conditions. Results regarding the relative strengths of these risk factors may not be generalizable outside of the Danish context. Our definition of urban residence was Denmark-specific. The definition of ADHD used here is reflective of a more severe hyperkinetic subtype of ADHD diagnosed by specialists, so that findings may not hold for a broader ADHD criterion commonly used in the US.

In conclusion, the study of etiologic heterogeneity is important when a diagnosis encompasses a variety of subtypes each resulting from a different profile of associated – potentially causal - factors. In this scenario, the search for causal factors for the single, overarching diagnosis group is hampered by ‘noise’ produced by varying effects among the subgroups. While some methods exist to assess etiologic heterogeneity, they are not applicable to diagnoses which can co-occur in the same person (non-competing events) when the probability of observing the presence or absence of co-occurring diagnoses accrue over time and the length of follow-up is variable. Such diagnostic co-occurrences are especially common for mental health conditions, and may also occur for cardiometabolic conditions. Our “subtraction” method reduces outcome misclassification, allows the inclusion of all persons in an open cohort, and provides an appropriate p value directly testing etiologic heterogeneity. This method may strengthen insight into diverse etiologic mechanisms that underlie phenotypic heterogeneity. Furthermore, this method has utility in studies using diagnostic data from large, population-based databases, data that is increasingly available and of high interest.

## Data Availability

The data in the present study comprise Danish registry data which, at the present time, can only be used by investigators residing in Denmark and collaborating with investigators of the National Center for Registry-based Research (NCRR) in Denmark.

https://github.com/jcyu46/X-Disorder-Code

