## Supplemental Tables for "Method to Test Etiologic Heterogeneity Among Non-Competing Diagnoses that Accrue Over Time: Application to Discriminating the Impact of Perinatal Exposures on Autism and Attention Deficit Hyperactivity Disorder"

Supplemental Digital Content

**eTable 1:** Numbers and Percents of Study Sample Who Would be Included and Excluded in Complete Case Analysis by Diagnostic Status

|  | Included^a^ | Excluded^b^ | Total |
| --- | --- | --- | --- |
|  | N=37305 | N=5526 | N=42831 |
| Sub-cohort | 16751 | 2494 (13.0%) | 19245 |
| Autism diagnosis | 11250^†^ | 1712 (13.2%) | 12962 |
| ADHD diagnosis | 12098^†^ | 1715 (12.4%)^†^ | 13813 |
| Autism + ADHD diagnoses | 2358 | 328 (12.2%) | 2686 |
| ^a^ Observations with no missingness for sex, birth year, parity, interpregnancy interval, maternal marital status, paternal age, maternal age, paternal and maternal history of psych diagnosis, paternal and maternal income level, paternal and maternal educational attainment,paternal and maternal employment category, paternal and maternal immigrant status, urbanicity, maternal smoking status | | | |
| ^b^ Observations with any missingness for any of the above variables | | | |

**dTable 2:** Characteristics of Persons Randomly Sampled from the Danish Population (iPSYCH Sub-cohort) Who Would be Included and Excluded in Complete Case Analysis

|  | Included^a^ | | Excluded^b^ | | Total | |
| --- | --- | --- | --- | --- | --- | --- |
|  | N=16751 | % | N=2494 | % | N=19245 | % |
| Sex |  |  |  |  |  |  |
| Female | 8169 | 48.8 | 1218 | 48.8 | 9387 | 48.8 |
| Male | 8582 | 51.2 | 1276 | 51.2 | 9858 | 51.2 |
| Birth year |  |  |  |  |  |  |
| 1991-1995 | 5568 | 33.2 | 1004 | 40.3 | 6572 | 34.1 |
| 1996-2000 | 5570 | 33.3 | 880 | 35.3 | 6450 | 33.5 |
| 2001-2005 | 5613 | 33.5 | 610 | 24.5 | 6223 | 32.3 |
| Parity |  |  |  |  |  |  |
| 1 | 7267 | 43.4 | 1220 | 48.9 | 8487 | 44.1 |
| 2 | 6430 | 38.4 | 756 | 30.3 | 7186 | 37.3 |
| > 3 | 3054 | 18.2 | 518 | 20.8 | 3572 | 18.6 |
| Interpregnancy interval |  |  |  |  |  |  |
| First birth | 7267 | 43,4 | 1220 | 48,9 | 8487 | 44,1 |
| < 2 years since prior birth | 1625 | 9,7 | 249 | 10,0 | 1874 | 9,7 |
| 2 - 4 years since prior birth | 4637 | 27,7 | 512 | 20,5 | 5149 | 26,8 |
| > 4 years since prior birth | 3222 | 19,2 | 513 | 20,6 | 3735 | 19,4 |
| Maternal marital status |  |  |  |  |  |  |
| Unmarried | 7982 | 47.7 | 900 | 36.1 | 8882 | 46.2 |
| Married | 8769 | 52.3 | 1594 | 63.9 | 10363 | 53.8 |
| Paternal age |  |  |  |  |  |  |
| Missing | - | - | 81 | 3.2 | 81 | 0.4 |
| < 20 years old | 193 | 1.2 | 33 | 1.3 | 226 | 1.2 |
| 21-25 years old | 1759 | 10.5 | 296 | 11.9 | 2055 | 10.7 |
| 26-30 years old | 5628 | 33.6 | 704 | 28.2 | 6332 | 32.9 |
| 31-35 years old | 5490 | 32.8 | 710 | 28.5 | 6200 | 32.2 |
| > 35 years old | 3681 | 22 | 670 | 26.9 | 4351 | 22.6 |
| Maternal age |  |  |  |  |  |  |
| < 20 years old | 551 | 3.3 | 135 | 5.4 | 686 | 3.6 |
| 21-25 years old | 3031 | 18.1 | 611 | 24.5 | 3642 | 18.9 |
| 26-30 years old | 6861 | 41 | 889 | 35.6 | 7750 | 40.3 |
| 31-35 years old | 4710 | 28.1 | 613 | 24.6 | 5323 | 27.7 |
| > 35 years old | 1598 | 9.5 | 246 | 9.9 | 1844 | 9.6 |
| Paternal Psychiatric Illness |  |  |  |  |  |  |
| No psychiatric diagnosis before index birth | 16380 | 97,8 | 2397 | 96,1 | 18777 | 97,6 |
| Psychiatric diagnosis before index birth | 371 | 2,2 | 97 | 3,9 | 468 | 2,4 |
| Maternal Psychiatric Illness |  |  |  |  |  |  |
| No psychiatric diagnosis before index birth | 16194 | 96,7 | 2407 | 96,5 | 18601 | 96,7 |
| Psychiatric diagnosis before index birth | 557 | 3,3 | 87 | 3,5 | 644 | 3,3 |
| Parental Psychiatric Illness |  |  |  |  |  |  |
| No psychiatric diagnosis before index birth | 15879 | 94,8 | 2324 | 93,2 | 18203 | 94,6 |
| Psychiatric diagnosis before index birth | 872 | 5,2 | 170 | 6,8 | 1042 | 5,4 |
| Paternal income decile |  |  |  |  |  |  |
| Missing | - | - | 169 | 6.8 | 169 | 0.9 |
| 1st | 1336 | 8 | 421 | 16.9 | 1757 | 9.1 |
| 2nd | 1478 | 8.8 | 449 | 18 | 1927 | 10 |
| 3rd | 1671 | 10 | 289 | 11.6 | 1960 | 10.2 |
| 4th | 1743 | 10.4 | 223 | 8.9 | 1966 | 10.2 |
| 5th | 1754 | 10.5 | 169 | 6.8 | 1923 | 10 |
| 6th | 1784 | 10.7 | 169 | 6.8 | 1953 | 10.1 |
| 7th | 1804 | 10.8 | 147 | 5.9 | 1951 | 10.1 |
| 8th | 1754 | 10.5 | 151 | 6.1 | 1905 | 9.9 |
| 9th | 1805 | 10.8 | 128 | 5.1 | 1933 | 10 |
| 10th | 1622 | 9.7 | 179 | 7.2 | 1801 | 9.4 |
| Maternal income decile |  |  |  |  |  |  |
| Missing | - | - | 438 | 17,6 | 438 | 2,3 |
| 1st | 1264 | 7,5 | 505 | 20,2 | 1769 | 9,2 |
| 2nd | 1591 | 9,5 | 265 | 10,6 | 1856 | 9,6 |
| 3rd | 1729 | 10,3 | 233 | 9,3 | 1962 | 10,2 |
| 4th | 1743 | 10,4 | 175 | 7,0 | 1918 | 10,0 |
| 5th | 1777 | 10,6 | 172 | 6,9 | 1949 | 10,1 |
| 6th | 1731 | 10,3 | 142 | 5,7 | 1873 | 9,7 |
| 7th | 1749 | 10,4 | 137 | 5,5 | 1886 | 9,8 |
| 8th | 1714 | 10,2 | 147 | 5,9 | 1861 | 9,7 |
| 9th | 1748 | 10,4 | 143 | 5,7 | 1891 | 9,8 |
| 10th | 1705 | 10,2 | 137 | 5,5 | 1842 | 9,6 |
| Paternal education level |  |  |  |  |  |  |
| Missing | - | - | 835 | 33.5 | 835 | 4.3 |
| Primary school | 3866 | 23.1 | 480 | 19.2 | 4346 | 22.6 |
| Vocational education: general & business | 1298 | 7.7 | 155 | 6.2 | 1453 | 7.6 |
| Vocational education/training: professional internships | 7255 | 43.3 | 599 | 24 | 7854 | 40.8 |
| Upper secondary education: shorter-term bachelors equiv, profession-oriented masters | 2675 | 16 | 243 | 9.7 | 2918 | 15.2 |
| Upper secondary education: longer-term university-based masters, research or PhD | 1657 | 9.9 | 182 | 7.3 | 1839 | 9.6 |
| Maternal education level |  |  |  |  |  |  |
| Missing | - | - | 899 | 36 | 899 | 4.7 |
| Primary school | 4574 | 27.3 | 660 | 26.5 | 5234 | 27.2 |
| Vocational education: general & business | 2710 | 16.2 | 266 | 10.7 | 2976 | 15.5 |
| Vocational education: professional internships | 5410 | 32.3 | 370 | 14.8 | 5780 | 30 |
| Upper secondary education: shorter-term bachelors equiv, profession-oriented masters | 2966 | 17.7 | 216 | 8.7 | 3182 | 16.5 |
| Upper secondary education: longer-term university-based masters, research or PhD | 1091 | 6.5 | 83 | 3.3 | 1174 | 6.1 |
| Paternal employment category |  |  |  |  |  |  |
| Missing | - | - | 234 | 9.4 | 234 | 1.2 |
| Upper management | 3166 | 18.9 | 295 | 11.8 | 3461 | 18 |
| Professional and mid-level employees | 7280 | 43.5 | 616 | 24.7 | 7896 | 41 |
| Unskilled, self-employed, other | 4386 | 26.2 | 642 | 25.7 | 5028 | 26.1 |
| On leave | 148 | 0.9 | 43 | 1.7 | 191 | 1 |
| Unemployed | 863 | 5.2 | 228 | 9.1 | 1091 | 5.7 |
| Pursuing education | 233 | 1.4 | 61 | 2.4 | 294 | 1.5 |
| Outside of labor force | 675 | 4 | 375 | 15 | 1050 | 5.5 |
| Maternal employment category |  |  |  |  |  |  |
| Missing | - | - | 531 | 21.3 | 531 | 2.8 |
| Upper management | 2504 | 14.9 | 219 | 8.8 | 2723 | 14.1 |
| Professional and mid-level employees | 7305 | 43.6 | 534 | 21.4 | 7839 | 40.7 |
| Unskilled, self-employed, other | 2817 | 16.8 | 323 | 13 | 3140 | 16.3 |
| On leave | 556 | 3.3 | 51 | 2 | 607 | 3.2 |
| Unemployed | 1379 | 8.2 | 210 | 8.4 | 1589 | 8.3 |
| Pursuing education | 745 | 4.4 | 113 | 4.5 | 858 | 4.5 |
| Outside of labor force | 1445 | 8.6 | 513 | 20.6 | 1958 | 10.2 |
| Paternal immigrant status |  |  |  |  |  |  |
| Missing | - | - | 81 | 3.2 | 81 | 0.4 |
| Not immigrant | 15240 | 91 | 1338 | 53.6 | 16578 | 86.1 |
| Immigrant | 1511 | 9 | 1075 | 43.1 | 2586 | 13.4 |
| Maternal immigrant status |  |  |  |  |  |  |
| Not immigrant | 15326 | 91.5 | 1374 | 55.1 | 16700 | 86.8 |
| Immigrant | 1425 | 8.5 | 1120 | 44.9 | 2545 | 13.2 |
| Urban Residence |  |  |  |  |  |  |
| Capital region | 2263 | 13.5 | 579 | 23.2 | 2842 | 14.8 |
| Suburb of capital | 2256 | 13.5 | 354 | 14.2 | 2610 | 13.6 |
| Near capital, having town with >10,000 population | 712 | 4.3 | 106 | 4.3 | 818 | 4.3 |
| Near capital, largest town with <10,000 population | 544 | 3.2 | 62 | 2.5 | 606 | 3.1 |
| Outside capital region, town with >100,000 population | 2087 | 12.5 | 330 | 13.2 | 2417 | 12.6 |
| Outside capital region, town with 40k-99,999 population | 1026 | 6.1 | 147 | 5.9 | 1173 | 6.1 |
| Outside capital region, town with 20k-39,999 population | 1778 | 10.6 | 216 | 8.7 | 1994 | 10.4 |
| Outside capital region, town with 10k-19,999 population | 1213 | 7.2 | 180 | 7.2 | 1393 | 7.2 |
| Other municipality, at least 50% live in urban area | 1256 | 7.5 | 144 | 5.8 | 1400 | 7.3 |
| Other municipality, <50% live in urban area | 3616 | 21.6 | 376 | 15.1 | 3992 | 20.7 |
| Maternal smoking status |  |  |  |  |  |  |
| Missing | - | - | 946 | 37.9 | 946 | 4.9 |
| Did not smoke during pregnancy | 12568 | 75 | 1230 | 49.3 | 13798 | 71.7 |
| Smoked during pregnancy | 4183 | 25 | 318 | 12.8 | 4501 | 23.4 |

^a^ Observations with no missingness for sex, birth year, parity, interpregnancy interval, maternal marital status, paternal age, maternal age, paternal and maternal history of psych diagnosis, paternal and maternal income level, paternal and maternal educational attainment,paternal and maternal employment category, paternal and maternal immigrant status, urban residence, maternal smoking status

^b^ Observations with any missingness for any of the above variables

**eTable 3**. Prevalence of Single and Joint Diagnoses of Autism and ADHD in 4 Closed Cohorts Constructed from the iPSYCH Study, Born Between 1991 and 2005, Denmark

|  | Subcohort | |  | Autism | | | | |  | ADHD | | | | |  | Autism+ADHD (Both) | | | | |
| --- | --- | --- | --- | --- | --- | --- | --- | --- | --- | --- | --- | --- | --- | --- | --- | --- | --- | --- | --- | --- |
| Closed Cohort | Actual | Weighted^a^ |  | Excl^b^ | Cont^c^ | New^d^ | Total | Prev |  | Excl^b^ | Cont^c^ | New^d^ | Total | Prev |  | Excl^b^ | Cont^c^ | New^d^ | Total | Prev |
| 6th birthday | 18768 | 938400 |  |  |  | 2807 | 2807 | 0.003 |  |  |  | 1064 | 1064 | 0.001 |  |  |  | 282 | 282 | 0.000 |
| 10th birthday | 14961 | 748050 |  | 821 | 1877 | 2529 | 4406 | 0.006 |  | 383 | 626 | 3409 | 4035 | 0.005 |  | 137 | 145 | 751 | 1060 | 0.001 |
| 14th birthday | 9984 | 499200 |  | 1924 | 2373 | 1854 | 4227 | 0.008 |  | 2236 | 1729 | 2008 | 3737 | 0.007 |  | 675 | 385 | 334 | 898 | 0.002 |
| 18th birthday | 4979 | 248950 |  | 2577 | 1591 | 1036 | 2627 | 0.011 |  | 2515 | 1183 | 1275 | 2458 | 0.010 |  | 636 | 262 | 117 | 477 | 0.002 |

a. Because the iPSYCH subcohort represented 2% of the total Danish birth population, we have constructed an approximate total cohort size by multiplying the actual included persons by 50.

b. Excl = excluded = persons who were included and diagnosed in the previous period, but no longer meet criteria for being included in this closed cohort. An example is a person diagnosed with autism who did not have sufficient follow up to be included.

c. Cont = continued = persons who were included and diagnosed with either a single or joint diagnosis and continued as being eligible and having the same diagnostic status in the next period.

e. New = persons who were newly diagnosed with a first or second diagnosis in this period.

**eTable 4**. Changes in Single and Joint Diagnoses of Autism and ADHD in 4 Closed Cohorts Constructed from the iPSYCH Study, Born Between 1991 and 2005, Denmark

|  | Autism | | | |  | ADHD | | | |  | Autism +ADHD | | | | | |
| --- | --- | --- | --- | --- | --- | --- | --- | --- | --- | --- | --- | --- | --- | --- | --- | --- |
| Closed Cohort | Cont ^a^ | New ^b^ | % 2^nd^ Dx ^c^ | Total |  | Cont ^a^ | New ^b^ | % 2^nd^ Dx ^c^ | Total |  | Autism to Both | ADHD to Both | New ^b^ | % 2^nd^ Dx ^d^ | % New ^d^ | Total |
| 6th birthday |  | 2807 |  | 2807 |  |  | 1064 |  | 1064 |  |  |  | 282 |  |  | 282 |
| 10th birthday | 1877 | 2529 | 5% | 4406 |  | 626 | 3409 | 8% | 4035 |  | 109 | 55 | 751 | 15% | 71% | 1060 |
| 14th birthday | 2373 | 1854 | 4% | 4227 |  | 1729 | 2008 | 4% | 3737 |  | 109 | 70 | 334 | 20% | 37% | 898 |
| 18th birthday | 1591 | 1036 | 4% | 2627 |  | 1183 | 1275 | 3% | 2458 |  | 59 | 39 | 117 | 21% | 25% | 477 |

a. Cont=continued. Persons diagnosed in the previous time period and continued as meeting cohort inclusions and not receiving a second diagnosis.

b. New = persons who were new to receiving a single or joint diagnosis in this time period.

c. % 2^nd^ Dx = the percent of persons with a single diagnosis who moved on to the next time period that received a second diagnosis.

d. % 2^nd^ Dx = the percent of those with joint diagnoses who received a second diagnosis only during this time period.

e. % New = the percent of those with joint diagnoses who received both the autism and ADHD diagnoses during this time period.

**eTable 5**. Comparison of Three Methods of Calculating Adjusted^a^ Measures of Association Between Urban Residence at Birth on Diagnosis of Autism and ADHD Separately and Jointly for iPSYCH Persons Born Between 1991 and 2005, Denmark

|  | Autism Only | | |  | ADHD Only | | |  | Autism+ADHD | | |
| --- | --- | --- | --- | --- | --- | --- | --- | --- | --- | --- | --- |
|  | RR | 95% CI | CLR ^b^ |  |  | 95% CI | CLR ^b^ |  |  | 95% CI | CLR ^b^ |
| **Closed Cohort Method** ^c^ | | | | | | | | | | | |
| 6th Birthday | 2.4 | 2.1, 2.7 | 1.3 |  | 1.7 | 1.3, 2.1 | 1.6 |  | 1.9 | 1.3, 2.9 | 2.3 |
| 10th Birthday | 1.8 | 1.6, 2.0 | 1.3 |  | 1.1 | 0.9, 1.2 | 1.3 |  | 1.9 | 1.5, 2.4 | 1.6 |
| 14th Birthday | 1.6 | 1.4, 1.9 | 1.3 |  | 1.1 | 0.9, 1.3 | 1.4 |  | 2.3 | 1.8, 2.9 | 1.7 |
| 18th Birthday | 1.5 | 1.3, 1.8 | 1.4 |  | 1.3 | 1.1, 1.6 | 1.5 |  | 2.3 | 1.7, 3.3 | 2.0 |
| **Open Cohort Assigned at End of Follow up** ^d^ | | | | | | | | | | | |
|  | 1.7 | 1.6, 1.8 | 1.2 |  | 1.1 | 1.1, 1.2 | 1.2 |  | 2.2 | 1.9, 2.5 | 1.3 |
| **New Subtraction Method** ^e^ | | | | | | | | | | | |
|  | 1.7 | 1.5, 1.9 | 1.2 |  | 1.2 | 1.1, 1.3 | 1.2 |  | 2.2 | 1.9, 2.5 | 1.3 |

a. All models were adjusted for sex, birth year, interpregnancy interval, maternal marital status, maternal and paternal age (quadratic term included), maternal and paternal psychiatric diagnosis prior to index birth, maternal and paternal income deciles (quadratic term included), maternal and paternal educational attainment, maternal and paternal employment status, maternal and paternal immigrant status, urbanicity and smoking.

b. CLR = confidence limit ratio, a measure of precision = 95% upper confidence limit/95% lower confidence limit.

c. A logistic regression model including persons who did not die or emigrate between birth and the 6^th^, 10^th^, 14^th^, or 18^th^ birthday and who were born in years allowing follow-up for the entire time frame (e.g. only persons born 1991-1994 had 18 years until the end of follow up in 2012).

d. A logistic regression model with the dependent variable of diagnosis assigned at the end of follow-up, which ranged from to 7 to 22 years.

e. Our primary analytic model, which calculates the risk of a single diagnosis (e.g. autism only) as the difference between the risk of being diagnosed with autism (autism only or autism+ADHD) and the risk of being diagnosed with autism+ADHD.

R Code Github Link

<https://github.com/jcyu46/X-Disorder-Code>

R Code

library(survival)

library(ggplot2)

dx1.time <- dx2.time <- comorbid.time <- seq(from=78, to=1144, by=52)

\# ---- Estimating ----

fit.dx1 = coxph(Surv(time = dx1_follow_duration, event = dx1) ~ exposure + confounders,

weights = weight, data = df)

fit.dx2 = coxph(Surv(time = dx2_follow_duration, event = dx2) ~ exposure + confounders,

weights = weight, data = df)

fit.comorbid = coxph(Surv(time = comorbid_follow_duration, event = comorbid) ~ exposure + confounders,

weights = weight, data = df)

\# Create high and low exposure dataframe:

predict.high <- predict.low <- df

predict.high$exposure = 1

predict.low$exposure = 0

\# In each group the base cumulative hazard is determined and saved as a function of the time:

tmp.dx1 = basehaz(fit.dx1)

H0 = stepfun(tmp.dx1$time, c(0, tmp.dx1$hazard))

tmp.dx2 = basehaz(fit.dx2)

H1 = stepfun(tmp.dx2$time, c(0, tmp.dx2$hazard))

tmp.comorbid = basehaz(fit.comorbid)

H2 = stepfun(tmp.comorbid$time, c(0, tmp.comorbid$hazard))

\# This is saved as in a matrix to prepare for calculating the survival function at each time point

H0.Mat = matrix(-H0(dx1.time))

H1.Mat = matrix(-H1(dx2.time))

H2.Mat = matrix(-H2(comorbid.time))

\# In each group the hazard is calculated for the high and low exposure data

\# Then the probability of having been diagnosed prior to each time point is calculated for both exposures

tmp.dx1.high = predict(fit.dx1, newdata = predict.high, type='risk')

tmp.dx1.low = predict(fit.dx1, newdata = predict.low, type='risk')

org.dx1.high = 1 - rowMeans(exp(H0.Mat %*% t(tmp.dx1.high)), na.rm = T)

org.dx1.low = 1 - rowMeans(exp(H0.Mat %*% t(tmp.dx1.low)), na.rm = T)

tmp.dx2.high = predict(fit.dx2, newdata = predict.high, type='risk')

tmp.dx2.low = predict(fit.dx2, newdata = predict.low, type='risk')

org.dx2.high = 1 - rowMeans(exp(H0.Mat %*% t(tmp.dx2.high)), na.rm = T)

org.dx2.low = 1 - rowMeans(exp(H0.Mat %*% t(tmp.dx2.low)), na.rm = T)

tmp.comorbid.high = predict(fit.comorbid, newdata = predict.high, type='risk')

tmp.comorbid.low = predict(fit.comorbid, newdata = predict.low, type='risk')

org.comorbid.high = 1 - rowMeans(exp(H0.Mat %*% t(tmp.comorbid.high)), na.rm = T)

org.comorbid.low = 1 - rowMeans(exp(H0.Mat %*% t(tmp.comorbid.low)), na.rm = T)

\# Calculating the Risk Ratios:

org.lnrr_dx1 = org.dx1.high/org.dx1.low

org.dx1.high.comorbid = org.dx1.high - org.comorbid.high

org.dx1.low.comorbid = org.dx1.low - org.comorbid.low

org.lnrr_dx1_comorbid = org.dx1.high.comorbid/org.dx1.low.comorbid

org.lnrr_dx2 = org.dx2.high/org.dx2.low

org.dx2.high.comorbid = org.dx2.high - org.comorbid.high

org.dx2.low.comorbid = org.dx2.low - org.comorbid.low

org.lnrr_dx2_comorbid = org.dx2.high.comorbid/org.dx2.low.comorbid

org.lnrr_comorbid = org.comorbid.high/org.comorbid.low

\# Making Plots

tmp.plot = as.data.frame(cbind(dx1.time, org.lnrr_dx1_comorbid, org.lnrr_dx2_comorbid, org.lnrr_comorbid))

colnames(tmp.plot) = c('time', 'dx1_Only', 'dx2_Only', 'Comorbid')

org.tmp <- ggplot() +

geom_point(data=tmp.plot, aes(time, dx1_Only), color='blue') +

geom_line(data=tmp.plot, aes(time, dx1_Only), color='blue') +

geom_point(data=tmp.plot, aes(time, dx2_Only), color='green') +

geom_line(data=tmp.plot, aes(time, dx2_Only), color='green') +

geom_point(data=tmp.plot, aes(time, Comorbid), color='purple') +

geom_line(data=tmp.plot, aes(time, Comorbid), color='purple') +

geom_hline(yintercept = 1, color = 'red', size = 1.5) +

labs(title = 'Risk Ratio for Original HR') +

xlab('Follow Duration in Days') +

ylab('Risk Ratio')

\# ---- Bootstrap ----

B = 100

\# matrices to contain results:

CI_matrix_dx1_high <- CI_matrix_dx1_low <- matrix(data = NA, nrow = B, ncol = length(dx1.time))

CI_matrix_dx2_high <- CI_matrix_dx2_low <- matrix(data = NA, nrow = B, ncol = length(dx2.time))

CI_matrix_comorbid_high <- CI_matrix_comorbid_low <- matrix(data = NA, nrow = B, ncol = length(comorbid.time))

for (b in 1:B){

set.seed(1111+b)

\# Sampling the weights:

df.temp = as.data.frame(df)

df.temp$weightexp <- df.temp$weightbs <- NA

index_control = which(df.temp$dx1 == 0 & df.temp$dx2 == 0)

df.temp$weightexp[index_control] = rexp(length(index_control))

df.temp$weightexp[index_control] = df.temp$weightexp[index_control]/mean(df.temp$weightexp[index_control])

index_dx1 = which(df.temp$dx1 == 1 & df.temp$dx2 == 0)

df.temp$weightexp[index_dx1] = rexp(length(index_dx1))

df.temp$weightexp[index_dx1] = df.temp$weightexp[index_dx1]/mean(df.temp$weightexp[index_dx1])

index_dx2 = which(df.temp$dx1 == 0 & df.temp$dx2 == 1)

df.temp$weightexp[index_dx2] = rexp(length(index_dx2))

df.temp$weightexp[index_dx2] = df.temp$weightexp[index_dx2]/mean(df.temp$weightexp[index_dx2])

index_comorbid = which(df.temp$dx1 == 1 & df.temp$dx2 == 1)

df.temp$weightexp[index_comorbid] = rexp(length(index_comorbid))

df.temp$weightexp[index_comorbid] = df.temp$weightexp[index_comorbid]/mean(df.temp$weightexp[index_comorbid])

df.temp$weightbs = df.temp$weight * df.temp$weightexp

df.temp$weightbs = round(df.temp$weightbs*(10^9), 0) # rounding the weights like this makes little to no difference and makes the code much faster to run due to R not having to handle long/small floats

\# the remainder of the code in the for-loop is identical to other case:

\# Fitting models

fit.dx1 = coxph(Surv(time = dx1_follow_duration, event = dx1) ~ exposure + confounders,

weights = weightbs, data = df.temp)

fit.dx2 = coxph(Surv(time = dx2_follow_duration, event = dx2) ~ exposure + confounders,

weights = weightbs, data = df.temp)

fit.comorbid = coxph(Surv(time = comorbid_follow_duration, event = comorbid) ~ exposure + confounders,

weights = weightbs, data = df.temp)

\# Create high and low exposure dataframe:

predict.high <- predict.low <- df

predict.high$exposure = 1

predict.low$exposure = 0

\# In each group the base cumulative hazard is determined and saved as a function of the time:

tmp.dx1 = basehaz(fit.dx1)

H0 = stepfun(tmp.dx1$time, c(0, tmp.dx1$hazard))

tmp.dx2 = basehaz(fit.dx2)

H1 = stepfun(tmp.dx2$time, c(0, tmp.dx2$hazard))

tmp.comorbid = basehaz(fit.comorbid)

H2 = stepfun(tmp.comorbid$time, c(0, tmp.comorbid$hazard))

\# This is saved as in a matrix to prepare for calculating the survival function at each time point

H0.Mat = matrix(-H0(dx1.time))

H1.Mat = matrix(-H1(dx2.time))

H2.Mat = matrix(-H2(comorbid.time))

\# In each group the hazard is calculated for the high and low exposure data

\# Then the probability of having been diagnosed prior to each time point is calculated for both exposures

tmp.dx1.high = predict(fit.dx1, newdata = predict.high, type='risk')

tmp.dx1.low = predict(fit.dx1, newdata = predict.low, type='risk')

CI_matrix_dx1_high[b,] = 1 - rowMeans(exp(H0.Mat %*% t(tmp.dx1.high)), na.rm = T)

CI_matrix_dx1_low[b,] = 1 - rowMeans(exp(H0.Mat %*% t(tmp.dx1.low)), na.rm = T)

tmp.dx2.high = predict(fit.dx2, newdata = predict.high, type='risk')

tmp.dx2.low = predict(fit.dx2, newdata = predict.low, type='risk')

CI_matrix_dx2_high[b,] = 1 - rowMeans(exp(H0.Mat %*% t(tmp.dx2.high)), na.rm = T)

CI_matrix_dx2_low[b,] = 1 - rowMeans(exp(H0.Mat %*% t(tmp.dx2.low)), na.rm = T)

tmp.comorbid.high = predict(fit.comorbid, newdata = predict.high, type='risk')

tmp.comorbid.low = predict(fit.comorbid, newdata = predict.low, type='risk')

CI_matrix_comorbid_high[b,] = 1 - rowMeans(exp(H0.Mat %*% t(tmp.comorbid.high)), na.rm = T)

CI_matrix_comorbid_low[b,] = 1 - rowMeans(exp(H0.Mat %*% t(tmp.comorbid.low)), na.rm = T)

}

\# Calculating the Risk Ratios:

lnrr_dx1_comorbid = (CI_matrix_dx1_high - CI_matrix_comorbid_high)/(CI_matrix_dx1_low - CI_matrix_comorbid_low)

sd_dx1_comorbid = apply(log(lnrr_dx1_comorbid), 2, sd, na.rm=T)

CI_U_dx1_comorbid = log(lnrr_dx1_comorbid) + qnorm(.975) * sd_dx1_comorbid

CI_L_dx1_comorbid = log(lnrr_dx1_comorbid) + qnorm(.025) * sd_dx1_comorbid

plot_dx1_comorbid = as.data.frame(cbind(dx1.time, lnrr_dx1_comorbid, exp(CI_U_dx1_comorbid), exp(CI_L_dx1_comorbid)))

colnames(plot_dx1_comorbid) = c('dx1.time', 'RR', 'RR_Upper', 'RR_Lower')

lnrr_dx2_comorbid = (CI_matrix_dx2_high - CI_matrix_comorbid_high)/(CI_matrix_dx2_low - CI_matrix_comorbid_low)

sd_dx2_comorbid = apply(log(lnrr_dx2_comorbid), 2, sd, na.rm=T)

CI_U_dx2_comorbid = log(lnrr_dx2_comorbid) + qnorm(.975) * sd_dx2_comorbid

CI_L_dx2_comorbid = log(lnrr_dx2_comorbid) + qnorm(.025) * sd_dx2_comorbid

plot_dx2_comorbid = as.data.frame(cbind(dx2.time, lnrr_dx2_comorbid, exp(CI_U_dx2_comorbid), exp(CI_L_dx2_comorbid)))

colnames(plot_dx2_comorbid) = c('dx2.time', 'RR', 'RR_Upper', 'RR_Lower')

lnrr_comorbid = (CI_matrix_comorbid_high)/(CI_matrix_comorbid_low)

sd_comorbid = apply(log(lnrr_comorbid), 2, sd, na.rm=T)

CI_U_comorbid = log(lnrr_comorbid) + qnorm(.975) * sd_comorbid

CI_L_comorbid = log(lnrr_comorbid) + qnorm(.025) * sd_comorbid

plot_comorbid = as.data.frame(cbind(comorbid.time, lnrr_comorbid, exp(CI_U_comorbid), exp(CI_L_comorbid)))

colnames(plot_dx1_comorbid) = c('comorbid.time', 'RR', 'RR_Upper', 'RR_Lower')

\# Making Plots

plot_all = as.data.frame(cbind(plot_dx1_comorbid, plot_dx2_comorbid, plot_comorbid))

plot_all = as.data.frame(plot_all[,-c(5,9)]) # remove duplicates of time

colnames(plot_all) = c('time',

'dx1.comorbid.RR', 'dx1.comorbid.RR.U', 'dx1.comorbid.RR.L',

'dx2.comorbid.RR', 'dx2.comorbid.RR.U', 'dx2.comorbid.RR.L',

'comorbid.RR', 'comorbid.RR.U', 'comorbid.RR.L')

p.all <- ggplot() +

geom_line(data=plot_all, aes(x = time, y=dx1.comorbid.RR, color='dx1')) +

geom_line(data=plot_all, aes(x = time, y=dx2.comorbid.RR, color='dx2')) +

geom_line(data=plot_all, aes(x = time, y=comorbid.RR, color='comorbid')) +

scale_colour_manual(name = 'Condition', values=c('dx1=blue', 'dx2'='green', 'comorbid=purple'),

labels = c('dx1', 'dx2', 'dx1+dx2')) +

ylab('Risk Ratio and 95% Confidence Interval') +

coord_trans(y='log10') +

scale_x_continuous(name = 'Follow-up Time (Years)') +

theme(axis.text.x = element_text(size=12, face='bold', angle=75)) +

ggtitle('Risk Ratio and 95% Confidence Interval for Exposure') +

geom_hline(yintercept = 1, color = 'red', size=1.5)

p.all <- p.all + geom_point(data=plot_all, aes(x=time, y=dx1.comorbid.RR), color='blue') +

geom_point(data=plot_all, aes(x=time, y=dx2.comorbid.RR), color='green') +

geom_point(data=plot_all, aes(x=time, y=comorbid.RR), color='purple') +

geom_ribbon(data=plot_all, aes(x=time, ymin=dx1.comorbid.RR.L, ymax=dx1.comorbid.RR.U), alpha=0.3, fill='blue') +

geom_ribbon(data=plot_all, aes(x=time, ymin=dx2.comorbid.RR.L, ymax=dx2.comorbid.RR.U), alpha=0.3, fill='green') +

geom_ribbon(data=plot_all, aes(x=time, ymin=comorbid.RR.L, ymax=comorbid.RR.U), alpha=0.3, fill='purple')

\# ---- p-values: average and whole curve ----

avgrr_dx1 = rowMeans(log(lnrr_dx1_comorbid))

avgrr_dx2 = rowMeans(log(lnrr_dx2_comorbid))

avgrr_comorbid = rowMeans(log(CI_matrix_comorbid_high/CI_matrix_comorbid_low))

org.avgrr_dx1 = mean(log(org.lnrr_dx1_comorbid))

org.avgrr_dx2 = mean(log(org.lnrr_dx2_comorbid))

org.avgrr_comorbid = mean(log(org.lnrr_comorbid))

p_vals = data.frame(matrix(NA, 2, 4))

colnames(p_vals) = c('all', 'dx1 vs dx2', 'dx1 vs comorbid', 'dx2 vs comorbid')

rownames(p_vals) = c('avg eq', 'whole curve eq')

\# average equal:

Sigmahat = var(cbind(avgrr_dx1, avgrr_dx2, avgrr_comorbid))

Lmat = rbind(c(1,-1,0),

c(0,1,-1))

est = Lmat %*% c(org.avgrr_dx1, org.avgrr_dx2, org.avgrr_comorbid)

estV = est %*% Sigmahat %*% est

chisq = t(est) %*% solve(estV) %*% est

pval = 1 - pchisq(chisq, df = Matrix::rankMatrix(estV))

p_vals_urban['avg eq', 'all'] = pval

Lmat = rbind(c(1,-1,0))

est = Lmat %*% c(org.avgrr_dx1, org.avgrr_dx2, org.avgrr_comorbid)

estV = est %*% Sigmahat %*% est

chisq = t(est) %*% solve(estV) %*% est

pval = 1 - pchisq(chisq, df = Matrix::rankMatrix(estV))

p_vals_urban['avg eq', 'dx1 vs dx2'] = pval

Lmat = rbind(c(1,0,-1))

est = Lmat %*% c(org.avgrr_dx1, org.avgrr_dx2, org.avgrr_comorbid)

estV = est %*% Sigmahat %*% est

chisq = t(est) %*% solve(estV) %*% est

pval = 1 - pchisq(chisq, df = Matrix::rankMatrix(estV))

p_vals_urban['avg eq', 'dx1 vs comorbid'] = pval

Lmat = rbind(c(0,1,-1))

est = Lmat %*% c(org.avgrr_dx1, org.avgrr_dx2, org.avgrr_comorbid)

estV = est %*% Sigmahat %*% est

chisq = t(est) %*% solve(estV) %*% est

pval = 1 - pchisq(chisq, df = Matrix::rankMatrix(estV))

p_vals_urban['avg eq', 'dx2 vs comorbid'] = pval

\# whole curve equal

Sigmahat = var(cbind(log(lnrr_dx1_comorbid)[,3:20],

log(lnrr_dx2_comorbid)[,3:20],

log(CI_matrix_comorbid_high/CI_matrix_comorbid_low)[,3:20]))

Lmat = cbind(diag(18), -diag(18), diag(rep(0, 18)))

est = Lmat %*% c(log(org.lnrr_dx1_comorbid)[3:20], log(org.lnrr_dx2_comorbid)[3:20], log(org.lnrr_comorbid)[3:20])

estV = est %*% Sigmahat %*% est

chisq = t(est) %*% ginv(estV) %*% est

pval = 1 - pchisq(chisq, df = Matrix::rankMatrix(estV))

p_vals_urban['whole cure eq', 'dx1 vs dx2'] = pval

Lmat = cbind(diag(18), diag(rep(0, 18)), -diag(18))

est = Lmat %*% c(log(org.lnrr_dx1_comorbid)[3:20], log(org.lnrr_dx2_comorbid)[3:20], log(org.lnrr_comorbid)[3:20])

estV = est %*% Sigmahat %*% est

chisq = t(est) %*% ginv(estV) %*% est

pval = 1 - pchisq(chisq, df = Matrix::rankMatrix(estV))

p_vals_urban['whole cure eq', 'dx1 vs comorbid'] = pval

Lmat = cbind(diag(rep(0, 18)), diag(18), -diag(18))

est = Lmat %*% c(log(org.lnrr_dx1_comorbid)[3:20], log(org.lnrr_dx2_comorbid)[3:20], log(org.lnrr_comorbid)[3:20])

estV = est %*% Sigmahat %*% est

chisq = t(est) %*% ginv(estV) %*% est

pval = 1 - pchisq(chisq, df = Matrix::rankMatrix(estV))

p_vals_urban['whole cure eq', 'dx2 vs comorbid'] = pval

Lmat = rbind(cbind(diag(rep(0, 18)), diag(18), -diag(18)),

cbind( diag(18), diag(rep(0, 18)), -diag(18)))

est = Lmat %*% c(log(org.lnrr_dx1_comorbid)[3:20], log(org.lnrr_dx2_comorbid)[3:20], log(org.lnrr_comorbid)[3:20])

estV = est %*% Sigmahat %*% est

chisq = t(est) %*% ginv(estV) %*% est

pval = 1 - pchisq(chisq, df = Matrix::rankMatrix(estV))

p_vals_urban['whole cure eq', 'all'] = pval
